# Reddit posts reveal how natural environments affect social anxiety in young people

**DOI:** 10.64898/2025.12.10.25341922

**Authors:** Karen O’Connor, Sophia Hernandez, Ana Lucia Schmidt, Patrick D. Shirey, Graciela Gonzalez-Hernandez

## Abstract

Social anxiety disorder affects over one-third of young people globally and is characterized by intense fear of social situations and concern about judgment from others. While effective treatments exist, many individuals remain undiagnosed or delay seeking treatment, highlighting the importance of identifying complementary strategies. Natural environments have shown benefit for mental health, yet their therapeutic potential for social anxiety remains understudied from the patient’s perspective.

We analyzed Reddit posts from adolescents and young adults in a social anxiety community to explore their experiences with green and blue spaces. From over 535,000 posts, we identified 1,887 nature-related posts through computational and manual review. The results showed a complex relationship between natural environments and social anxiety symptoms. While many users reported that being in the outdoors and outdoor activities helped reduce their anxiety and improved their moods, others described the environments as triggering due to fears of being observed or judged.

Our findings suggest that natural environments may be perceived as helpful, accessible options for managing social anxiety, particularly when combined with physical activity. However, interventions must be carefully designed to address the unique social evaluation concerns that distinguish social anxiety from other anxiety disorders. These insights could inform the development of nature-based approaches that complement traditional treatments.

## 1. Introduction

Anxiety is a normal response to stressors; however, when anxiety persists and disrupts daily living it may be classified as a disorder ^1^. Among the various anxiety disorders identified in the DSM-5 ^2^, social anxiety disorder (SAD) is particularly prevalent mental health condition, affecting an estimated 36% of young people globally ^3^. SAD differs from generalized anxiety as it is characterized by an intense fear of social situations and worry about being judged or scrutinized by others ^4^. Symptoms of social anxiety include difficulty being around unfamiliar people, becoming speechless, embarrassment, blushing, sweating, panic, and difficulty talking to people ^5^.

The population most at risk of social anxiety is adolescents and young adults between the ages of 18 and 24 ^3^, with the onset of symptoms occurring by age 11 in 50% of cases, and by age 20 in 80% of cases ^6^. This age of onset coincides with major developmental periods in life, potentially impacting social, educational, and occupational development ^7^. Indeed, studies have shown the severity and profound impact that SAD has on young adults. SAD significantly impairs educational achievement and persistence, work performance, and social relationships ^8,9^. A large-scale study found that 92.6% of individuals with SAD reported significant interference in their daily lives ^3^. When compared to those with major depressive disorder, individuals with SAD report having a lower quality of life ^10^.

Evidence-based treatment options are available for alleviating social anxiety symptoms, including psychotherapy approaches, such as cognitive behavioral therapy, or pharmacotherapy interventions, including antidepressants or anti-anxiety medications ^11,12^. While effective treatments for SAD exist, they typically require professional intervention. However, many individuals with social anxiety remain undiagnosed or do not seek help promptly ^13,14^. Very likely due to the nature of the disorder, studies indicate that only 15-25% ^15,16^of individuals with SAD ever seek treatment, and the median delay between onset and first treatment contact is 16 years ^15^. Thus, exploring and quantifying the impact of alternative or complementary strategies that are reported to alleviate symptoms and improve the quality of life for individuals with SAD is important. In this study, we explore the potential therapeutic benefit of exposure to natural environments on SAD, particularly green and blue spaces, directly from patient reports.

Green spaces (areas of land that contain vegetation such as parks, forests, and gardens ^17^) and blue spaces (outdoor spaces that prominently feature water, including lakes, rivers, and coastal areas) have been associated with many positive benefits to health and well-being in the general population ^18,19^. These natural environments have been linked to reduced stress, improved mood, enhanced cognitive function, and improved mental health ^19,20^. Studies focusing specifically on SAD and its relation to green and blue spaces are limited, particularly for young people. A recent review found 54 studies focusing on depression and anxiety that used Reddit as their data source ^21^. Most studies were focused on depression, and many used the data to create prediction models. None of the anxiety studies explicitly included SAD, or social phobia, in their analysis. To the best of our knowledge, no study to date has exclusively examined patients’ reported experience with the natural environment and its effects on their SAD. The specific impact of these spaces on individuals with SAD remains understudied, particularly from the patient perspective.

Several theoretical frameworks may help explain the potential therapeutic benefits of natural environments for SAD. Attention Restoration Theory (ART) ^22^ suggests that natural environments may help restore one’s ability to concentrate and focus. For individuals with SAD, whose cognitive resources may be depleted by hypervigilance and self-monitoring in social situations^23^, natural environments may help reduce mental fatigue and restore focus. Stress Reduction Theory (SRT) ^24^ postulates that exposure to natural environments triggers responses that reduce the physiological effects of stress and promote recovery. This may be relevant for SAD, where physical symptoms of social anxiety, such as increased heart rate, rapid breathing, and muscle tension, can occur. The Biophilia Hypothesis ^25^ proposes that humans tend to connect with nature and other living things. This framework may help explain why natural environments might feel less threatening than other social or artificial environments, as these environments may provide opportunities for connection and engagement with others within the calming effects of nature. However, while natural environments may offer therapeutic benefits, they may also present unique challenges for individuals with SAD, eliciting responses like those in artificial environments, such as fear of negative evaluation during outdoor activities or anxiety about unexpected social encounters in public spaces. How exactly individuals with SAD engage with these environments could inform the development of nature-based interventions specifically tailored to address social anxiety symptoms.

Our study aims to bridge these gaps in knowledge by extracting and analyzing the experiences and perceptions of individuals with SAD with respect to their interactions with green and blue spaces. By focusing on the patient’s perspective, we will gain insights into how these natural environments, or activities in these environments, are perceived to impact social anxiety symptoms and affect overall well-being. Patients with chronic conditions often turn to social media to interact with peers, looking to share experiences or get advice or support ^26,27^. Studies have shown that individuals with SAD prefer online over face-to-face interactions ^28^ and experience lower anxiety when interacting online ^29^, particularly if they can control how they present themselves and can provide some level of anonymity ^30^. With this in mind, our study conducts a content analysis of Reddit posts to understand the perspectives of individuals with SAD regarding their interactions with natural spaces, thus providing insight into the potential for therapeutic benefits of natural spaces for this hard-to-reach and understudied community. Reddit is a social forum website where users post and interact with one another based on topics of interest. These focused groups have their conversations in dedicated forums known as subreddits. Unlike many other social media platforms, Reddit allows users to post anonymously, an appealing feature for those with mental health disorders, as they feel less stigma and post with less inhibition ^31^.

## 2. Methods

### 2.1 Data Collection

Reddit was selected as the data source for this study due to the availability of the data, the self-identified cohort, and the conversational style of the posts, which provide rich, detailed accounts of personal experiences. All posts from the r/*socialanxiety* subreddit were retrieved using the Pushshift API on March 23, 2022. Pushshift was a social media data collection, analysis, and archiving platform that collected Reddit data and made it available to researchers from 2015 until 2023 ^32^. While the Pushshift API is no longer available, the data can still be found at Academic Torrents ^33^.

The dataset includes all the posts on the r/*socialanxiety* subreddit (n = 535,544) available at the download date. The data collected is composed of post ID, author, title of the thread (if it’s the original post in the thread), post text, and timestamp.

### 2.2 Age Filtering

The data was then processed to detect users who reported being between 13 and 25 years old at the time of writing the posts ^34^. To do this, we first cleaned the posts by removing all URLs and replacing written numbers in the 1 to 99 range with their equivalent 2-digit representation. Secondly, we removed all posts that do not have numbers in the 13 to 25 range. Thirdly, we use regular expressions to automatically detect sentences that probably do not contain the self-reported age of the user. Some of these regular expressions identify numbers that refer to periods (hours, minutes, seconds, days, etc.), to dosage (pill, capsule, vial, drops, etc.), and to other common expressions that use 2-digit numbers that are clearly not related to age (catch-22, 24/7, money, percentages, COVID-19, etc.). Finally, the remaining 14,027 posts (from 10,670 users) and their corresponding sentences were annotated. The annotator labeled the posts with the reported age on the text or NA if no age could be determined. Given that the age detection was done on posts, it’s possible for a user to have multiple reported ages throughout their time on r/*socialanxiety*. Thus, we estimated the final age of each user by first calculating their birth year and then calculating the median birth year of each user. Therefore, the estimated age of the user will be their age in the most active year within the subreddit, that is, the most active year minus birth year. Given that the user activity in days has a long tail distribution where median=16 days, mean=181 days, and sd=322 days, we believe it is a fair approximation of the user’s age.

### 2.3 Keyword filtering

From 535,544 posts in the *r/socialanxiety* subreddit, there were 101,704 posts from 7,645 users classified as aged 13-25. These 101,704 posts were filtered using a comprehensive list of outdoor-pertinent keywords related to green and blue spaces that we developed. To identify keywords related to outdoor spaces and activities, we first tokenized the corpus and reviewed terms that occurred more than once in the corpus. The curated list of words included green spaces (e.g., park, forest, garden), blue spaces (e.g., beach, ocean, lake), as well as outdoor activities (e.g., running, hiking, swimming). A total of 108 keywords were identified, 75 of which were related to green spaces, with 43 potentially referencing natural spaces and 32 potentially referencing outdoor activities. The remaining 33 keywords were related to blue spaces, with 20 potentially referencing natural spaces and 13 potentially referencing outdoor activities. The list of keywords used can be found in Table 1. To reduce the number of posts for manual review, regular expressions were developed from a review of the keyword results to further filter the set by eliminating false-positive posts. The regular expressions filtered common expressions that would be captured with the keywords, such as “*running on empty*” or “*rock the boat*”. These also removed common cultural references, such as the television show “*Parks and Recreation*”. The total number of mentions in the filtered corpus is reported in Table 1. The complete regular expression patterns are in Supplemental Material (S1). Figure 1 shows an overview of the data selection process. The final dataset consisted of 4,070 posts from 2,082 users. All keyword and regular expression filtering were performed in RStudio version 2022.02.3+492.

**Figure 1:**
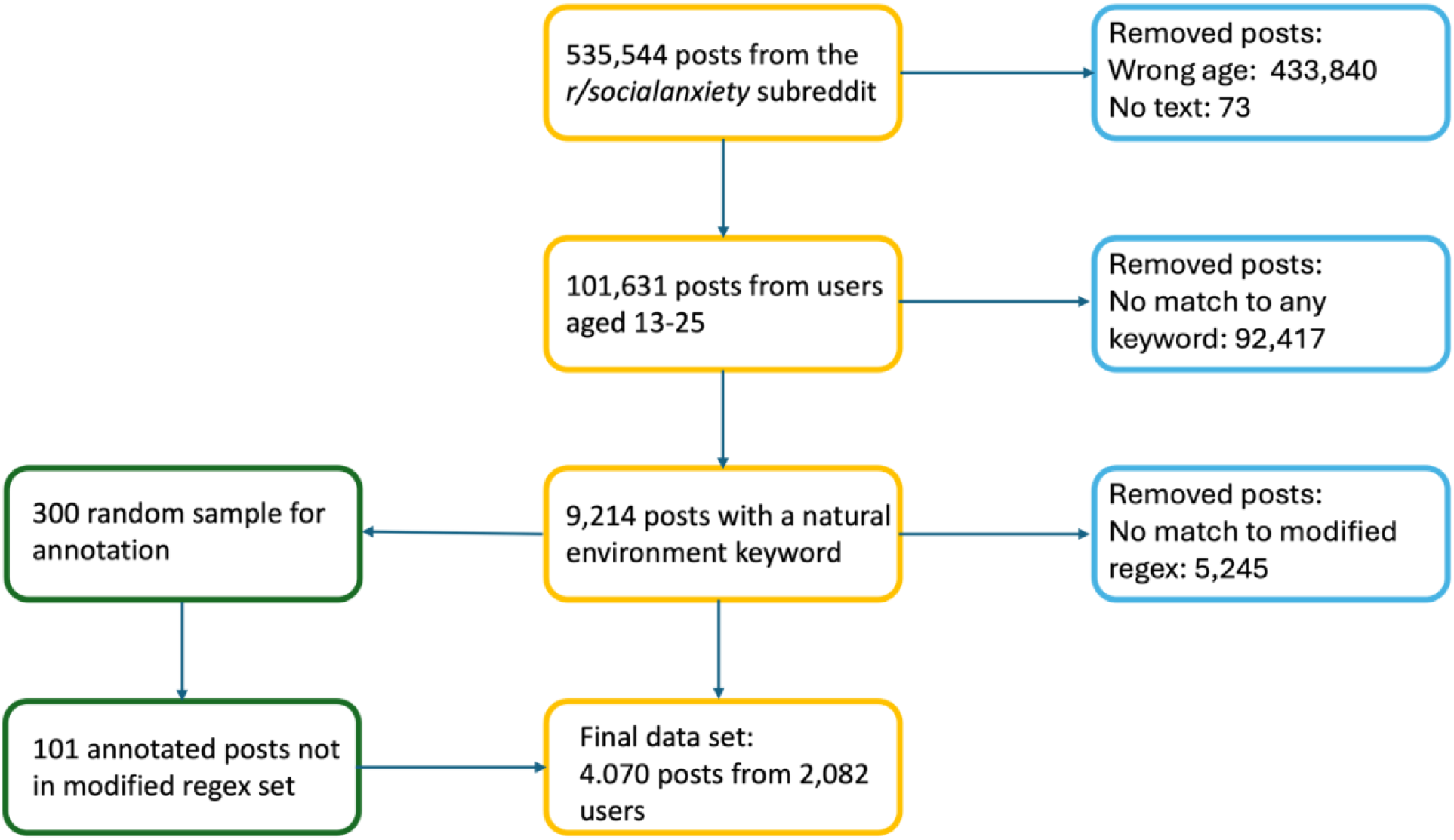
To create the data set for annotation, the original corpus was filtered first by the user’s age, then by natural environment keywords. The posts with keywords were further filtered using regular expressions to reduce the number of false positive results prior to manual review.

**Table 1:** Keywords identified as potentially referencing green or blue spaces, or outdoor activities in those spaces. The keyword count is the total number of matches for the term in the corpus, and the modified counts are the counts after more precise regular expressions were applied.

| Green Spaces Related Terms |  |  |  |  |  | Blue Spaces Related Terms |  |  |  |  |  |
| --- | --- | --- | --- | --- | --- | --- | --- | --- | --- | --- | --- |
| Place |  |  | Activity |  |  | Place |  |  | Activity |  |  |
| Keyword | Keyword<br>Regex<br>Count | Modified<br>Regex<br>Count | Keyword | Keyword<br>Regex<br>Count | Modified<br>Regex<br>Count | Keyword | Keyword<br>Regex<br>Count | Modified<br>Regex<br>Count | Keyword | Keyword<br>Regex<br>Count | Modified<br>Regex<br>Count |
| air | 180 | 42 | archery | 7 | 7 | bay | 22 | 2 | boat | 348 | 45 |
| back yard | 51 | 51 | backpacking | 11 | 11 | beach | 165 | 167 | canoe | 3 | 3 |
| ballpark | 4 | 4 | badminton | 3 | 3 | coast | 24 | 0 | cruise | 10 | 10 |
| barbecue | 32 | 32 | baseball | 33 | 29 | creek | 5 | 4 | dive | 75 | 75 |
| barn | 11 | 11 | basketball | 127 | 128 | dock | 9 | 9 | fishing | 23 | 23 |
| bench | 70 | 66 | bicycle | 234 | 236 | ferry | 2 | 2 | kayakers | 6 | 6 |
| bleachers | 12 | 12 | climb | 121 | 33 | harbor | 3 | 3 | lifeguard | 3 | 3 |
| bonfires | 13 | 13 | football | 140 | 140 | lake | 27 | 27 | paddleboard | 2 | 2 |
| breeze | 12 | 7 | frisbee | 4 | 4 | ocean | 22 | 22 | rowing | 10 | 6 |
| cabin | 22 | 22 | golf | 42 | 42 | pond | 4 | 4 | sail | 21 | 21 |
| camp | 137 | 116 | handball | 3 | 3 | pool | 116 | 109 | scuba | 5 | 5 |
| cave | 29 | 29 | hike | 110 | 111 | river | 19 | 15 | surf | 30 | 16 |
| cliff | 16 | 16 | horse | 53 | 52 | sand | 23 | 18 | swim | 155 | 155 |
| cookout | 6 | 6 | hunt | 88 | 49 | sea | 54 | 37 |  |  |  |
| countryside | 15 | 15 | jog | 73 | 73 | shore | 7 | 7 |  |  |  |
| courtyard | 7 | 7 | lacrosse | 13 | 13 | stream | 72 | 30 |  |  |  |
| daylight | 14 | 14 | long boarding | 3 | 3 | tide | 8 | 8 |  |  |  |
| desert | 13 | 13 | parkour | 6 | 6 | water | 364 | 9 |  |  |  |
| farm | 27 | 27 | quarterback | 2 | 2 | waterfall | 6 | 6 |  |  |  |
| footpath | 3 | 3 | riding | 91 | 69 | waves | 48 | 12 |  |  |  |
| forest | 27 | 27 | roller blade | 9 | 10 |  |  |  |  |  |  |
| fountain | 7 | 7 | rugby | 6 | 6 |  |  |  |  |  |  |
| garden | 59 | 53 | run | 1883 | 944 |  |  |  |  |  |  |
| grass | 30 | 14 | skate | 81 | 113 |  |  |  |  |  |  |
| hill | 31 | 14 | ski | 38 | 38 |  |  |  |  |  |  |
| landscape | 11 | 8 | snowboard | 14 | 14 |  |  |  |  |  |  |
| lawn | 18 | 18 | soccer | 123 | 127 |  |  |  |  |  |  |
| mountain | 91 | 74 | softball | 5 | 5 |  |  |  |  |  |  |
| nature | 347 | 133 | stroll | 7 | 7 |  |  |  |  |  |  |
| open space | 7 | 7 | tennis | 46 | 46 |  |  |  |  |  |  |
| outdoor | 70 | 70 | volleyball | 27 | 27 |  |  |  |  |  |  |
| outside | 2494 | 696 | walk | 4677 | 414 |  |  |  |  |  |  |
| park | 327 | 263 |  |  |  |  |  |  |  |  |  |
| patio | 3 | 3 |  |  |  |  |  |  |  |  |  |
| picnic | 21 | 21 |  |  |  |  |  |  |  |  |  |
| play ground | 26 | 26 |  |  |  |  |  |  |  |  |  |
| rink | 12 | 13 |  |  |  |  |  |  |  |  |  |
| sun | 170 | 170 |  |  |  |  |  |  |  |  |  |
| tree | 64 | 56 |  |  |  |  |  |  |  |  |  |
| vegetation | 2 | 2 |  |  |  |  |  |  |  |  |  |
| wilderness | 9 | 9 |  |  |  |  |  |  |  |  |  |
| wildlife | 9 | 3 |  |  |  |  |  |  |  |  |  |
| wind | 61 | 48 |  |  |  |  |  |  |  |  |  |

### 2.4 Annotation

Posts were manually coded in two rounds of annotation. Detailed annotation guidelines were developed to ensure consistent classification in both rounds of the annotation process. In the first round, posts were classified as “Nature-Related” or “Non-Nature Related.” Nature-related posts explicitly referenced natural environments, outdoor spaces, or outdoor activities. Non-Nature related posts used keywords in other contexts, such as metaphors (e.g., “waves of anxiety”), media references (e.g., “Forest Gump”), homonyms (e.g., “park the car”), or indoor activities (e.g., “rowing at the gym”). Examples of each classification are provided in Table 2.

**Table 2:** Definition for each annotation category, first by classification as ‘Nature’ or ‘Non-nature’ related. Nature-related posts were then classified as ‘Positive’, ‘Negative’, or ‘Neutral’ based on the annotation guidelines. Examples of posts are given for each category.

| Code | Definition | Example |
| --- | --- | --- |
| Round 1: Nature or Non_nature |  |  |
| Nature | The keyword is referencing a natural environment, being outside, or participating in an activity in a natural environment. | *"However **pools and beaches** always have a lot of people. It is terrifying for me to be partially undressed near so many strangers, it would make me unable to enjoy the water."* |
| Non_nature | The keyword is not related to nature, or the activity does not happen in a natural environment. | *"a foreign tourist stopped me, asking for help with a photograph. . After agreeing, I asked "this way?" (indicating **landscape or portrait mode**?)"* |
| Round 2: Impact on SAD |  |  |
| Positive | Positive posts are ones where the users mention that nature helped manage or alleviate SAD symptoms, including explicit statements of benefit, recommendations to others, or | *"One beneficial aspect of social anxiety I find is it motivates me to maintain my fitness (when I **run**, I feel a boost in serotonin that reduces my social anxiety symptoms for many hours) and make nutritious food choices. "* |
|  | actively seeking nature-based activities for symptom relief.<br>For example:<br>- If the user is recommending the environment or activity<br>- If the keyword is used to describe helping themselves or someone else |  |
| Negative | Posts should be coded as negative if they are describing nature as ineffective or exacerbating symptoms, including expressions of avoidance or negative experiences being in natural environments or with outdoor activities.<br>For example:<br>- They mention they don't, can't or won't like the nature-related keyword<br>- They mention the keyword in a negative way | *"I live near a **lake** but going by myself makes my anxiety symptoms worse, and I have no friends to go with. I'm afraid of running into former classmates, and feeling anxious and being alone does not seem like a good time."* |
| Neutral | Posts mentioning nature without clear connection to the user's SAD or any effects on it, such as general descriptions, hypothetical scenarios, or references to events that happened in the past without current relevance. | *"My state's cases exceed that of most nations. The lack of concern is unreal, amusement parks are not closing, beaches are full, people are eating out in restaurants. "* |

In the second round, Nature-Related posts were then sub-categorized based on the perceived impact of the natural environment or outdoor activity on users’ SAD as positive, negative, or neutral. We defined these categories as follows:

- Positive Impact: Posts indicating nature helped manage or alleviate SAD symptoms, including explicit statements of benefit, recommendations to others, or actively seeking nature-based activities for symptom relief.
- Negative Impact: Posts describing nature as ineffective or exacerbating symptoms, including expressions of avoidance or negative experiences from being in natural environments or with outdoor activities.
- Neutral Impact: Posts mentioning nature without an apparent connection to the user’s SAD or any effects on it, such as general descriptions, hypothetical scenarios, or references to events that happened in the past without current relevance.

The annotation was performed in an Excel spreadsheet that provided the annotators with the post metadata (post ID, user name, post date, etc.) and the title and text of the post. Annotators were also provided with the sentence in which the keyword(s) appeared in the post. This information was provided to facilitate the finding of the keyword in lengthy posts; however, the annotators’ decisions were based on information from the whole post. While a post may have multiple keywords matched, posts were limited to only one impact code. Posts with at least one positive impact related to a keyword were coded as positive. Neutral impact posts only contain mentions defined as neutral. Table 2 provides representative examples of posts in each impact category.

Annotation was conducted by two researchers who are experienced in annotation and trained on our annotation guidelines. Through an iterative process, each researcher independently annotated a portion of a random subset (N=1063) of the posts, after which disagreements were discussed and adjudicated through consensus. Any needed revisions to the guidelines were made after each iteration. This initial subset of posts ensured coverage of all keywords by randomly selecting posts from each keyword matched. After substantial inter-annotator agreement (IAA) was reached and no further revisions to the guidelines were deemed necessary, the last set (n = 613/1063) was double annotated, achieving an IAA of k = 0.80 for nature-related categorization and k=0.73 for the impact classification, which indicates strong and moderate agreement ^35^. Due to the high concordance reached, the remaining posts were independently annotated. Code and annotation guidelines are available at [link provided on publication].

### 2.5 Topic Modeling

We applied BERTopic to all posts classified as ‘Nature Related’ to identify latent topics of discussion within the corpus. BERTopic is a state-of-the-art topic modeling technique using the transformer model BERT and class-based TF-IDF ^36^.

Before applying the topic model, we performed several preprocessing steps. First, we identified identical or near-identical posts from the same user using cosine similarity. Posts with a similarity score > 0.90 were reviewed and removed as needed. Next, we preprocessed the data to clean and normalize the text. The following text preprocessing steps were taken: (1) mentions of social anxiety disorder and their variant (e.g., SA, social anxiety, SAD) were normalized to the term ‘*social_anxiety*’ and similarly, mentions of generalized anxiety disorder (e.g., GAD, GA, generalised anxiety) were normalized to ‘*generalized_anxiety*’; (2) all contractions were expanded; (3) exaggerations in text leading to misspellings (e.g., ‘loooong’, ‘saaaadddd’, etc.) were corrected; (4) all text was converted to lower case and punctuation was removed; and (5) posts were segmented into paragraphs. Paragraph segmentation was necessary to avoid the truncation of posts by the embedding model due to token limitations in the context window. The step also mitigates the issue of BERTopic only assigning one topic per document. By assigning a topic to each paragraph of the post, the results can be concatenated to identify multiple topics per post. Paragraph segmentation was done using the natural breaks in Reddit posts.

Using the BERTopic algorithm, the preprocessed text was passed through the sentence transformer model, all-mpnet-base-v2, to convert the text to numerical representations. This transformer model was fine-tuned on a diverse dataset of over 1.1 billion sentence pairs, including Reddit comments and academic texts. It is optimized for semantic similarity, making it well-suited for clustering text embeddings ^37^. Next, dimensionality reduction was performed using uniform manifold approximation and projections (UMAP). UMAP is a technique that keeps the data’s structure while reducing the dimensionality needed for clustering. The algorithm, Hierarchical Density-Based Spatial Clustering of Applications with Noise (HDBScan), was used for clustering. Finally, the c-TF-IDF algorithm was used to identify the representative word of the topic. In this algorithm, topic representation words are derived from the calculated clusters rather than individual documents.

The BERTopic topic model was fine-tuned for optimization through the customization of parameters for dimensionality reduction (UMAP), such as n_neighbors and n_components, clustering (HDBScan) parameters, such as cluster size and clustering selection methods, and topic representations (c-TF-IDF). The topic representations were optimized by removing stop words, the keywords used to filter the posts, and lemmatizing terms. The preprocessing and BERTopic model were implemented in Python v3.10. All code is available at: [link provided on publication].

### 2.5 Qualitative Analysis

We conducted a manual qualitative analysis to delve more deeply into themes specific to the natural environment keywords in the posts and the users’ perceived impact of these spaces or activities on their SAD. Posts that were identified as having a positive or negative impact were included in the analysis. Neutral posts were excluded as they, by definition, reported no impact on the user’s SAD.

A mixed-method, deductive and inductive, approach to coding was used to develop the codes and themes used in our analysis. To develop a deductive coding framework, we drew upon several relevant theoretical frameworks related to the natural environment and its proposed effects on mental health and well-being. These frameworks included Attention Restoration Theory (ART)^22^, which posits that natural environments can help restore depleted cognitive resources, Stress Reduction Theory (SRT)^24^, which proposes that exposure to natural environments can reduce stress, ease states of alert, and promote well-being, and the Biophilia hypothesis ^25^, which postulates that humans tend to want to connect with nature and other living things. We also allowed for inductive coding, where codes were added based on what was discovered in the data.

Our initial codebook was developed within the deductive framework using the familiarity with the data that was derived from the initial categorization of the data (described in Section 2.3 Keyword Filtering). Table 3 provides the codes and their definitions. Two researchers who had initially categorized the posts independently coded the data using NVivo v14. An initial set of 20 posts was coded to test the guidelines and identify any emergent codes from the data. Disagreements from the set were resolved by consensus, and the codebook was clarified with new codes added. After completing the set, a second set of 50 posts was coded with discussion and adjudication occurring between the two coders. Minimal guideline changes were made at this point, and coding was completed on the remaining set (n = 237). Upon completion, the disagreements were resolved. At this point, code saturation was determined to have occurred as no new codes were emerging from the data.

**Table 3:**
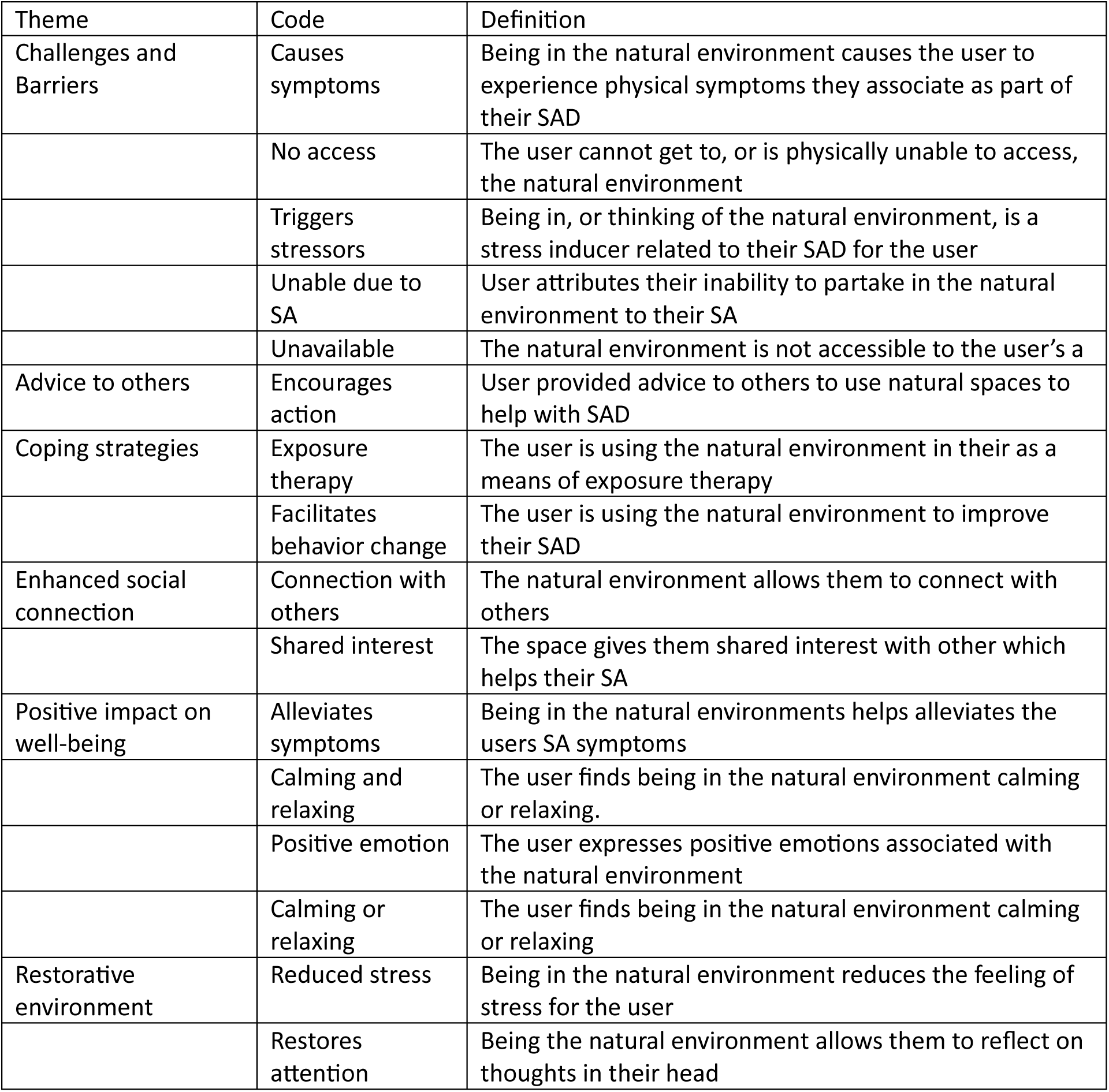
Codes and their definitions that were used for the analysis of the text. The resulting themes discovered in the data are also shown.

We identified 16 codes in the data. These were sorted into common themes prior to analysis. We identified six overarching themes in the data (Table 3).

## 3. Results

### 3.1 Annotation

In total, 4,070 posts were manually categorized as nature-related or not nature-related. 2,159 were determined to be non-nature, and 1,899 were nature. Before analysis, the nature posts were further filtered using cosine similarity to remove similar posts from the same users. After identifying and reviewing semantically similar texts, 12 posts were removed, resulting in 1,887 nature posts remaining for analysis. Posts determined as nature were sorted by impact. 544 were coded as positive, 272 were coded as negative, and 1071 were coded as neutral.

In total, 5,254 unique-per-post keywords were mentioned in the 4,046 included posts (Figure 2). The majority of these mentions were related to green spaces, with 2,447 mentions of green activity (46.6%) and 1,992 (37.9%) mentions of green places. Blue activity had only 295 mentions (5.6%), and blue place keywords had 520 mentions (9.9%). The total keyword mentions were somewhat equally divided between nature (n=2,842, 54.1%) and non-nature (n=2,412, 45.9%) classification.

**Figure 2:**
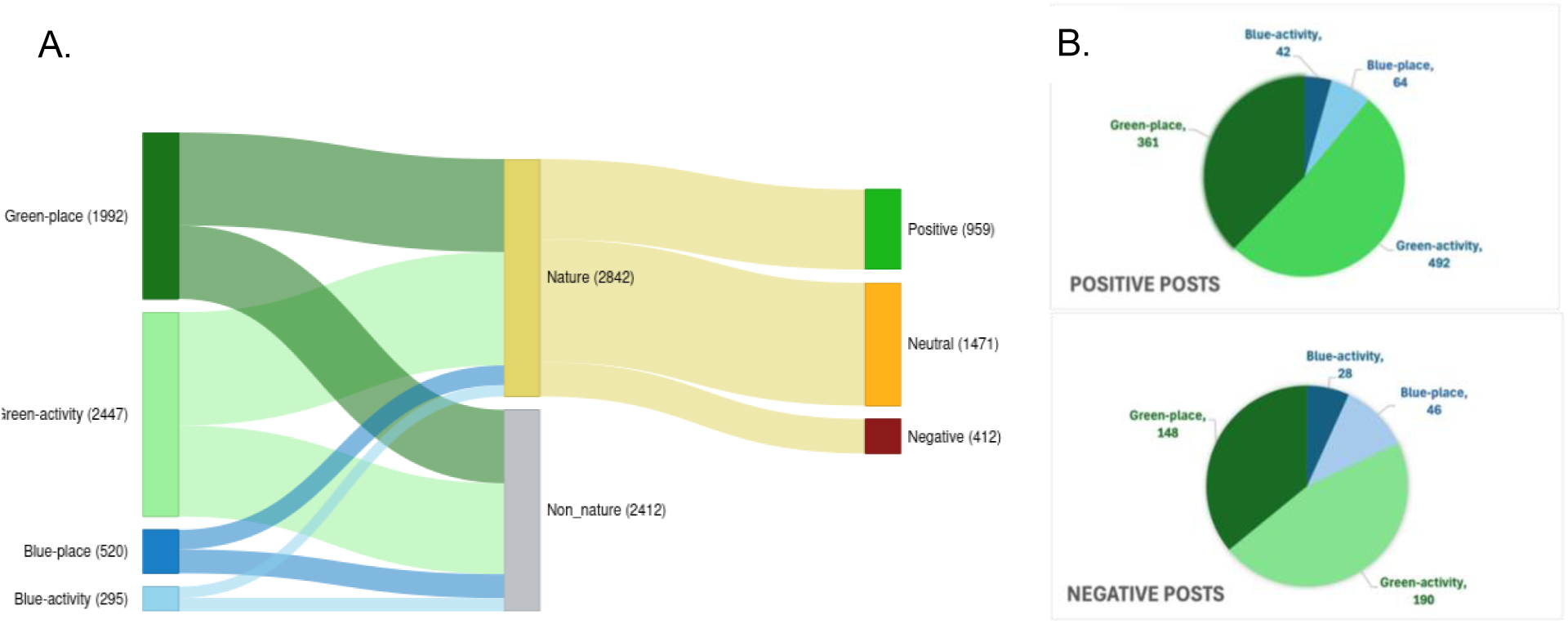
A. Sangkey chart representing the type of keyword mentions in posts, showing the breakdown of their nature-related classification and the nature-related impact classification. B. Keyword counts by environment type for positive and negative posts.

The majority of the keyword mentions were in posts classified as either non-nature related or neutral mentions (n= 3,883, 73.9%). The top 5 keywords mentioned for each group, with their classification counts, are shown in Table 4. Within the posts classified as positive, there were 831 keywords related to the green environment, with most (59.2%, 492/831) related to activities. For blue environment keywords, there were more keywords related to places (60.3%, 64/106). The negative posts followed a similar pattern with green environment activity-related keywords (56.2%, 190/338) and blue environment place keywords (62.2% 46/74) predominant.

**Table 4:** Top 5 keywords by total count of unique-per-post mentions in the corpus by keyword subtype.

| Keyword | Nature_positive | Nature_neutral | Nature_negative | Non_nature | Total | Keyword | Nature_positive | Nature_neutral | Nature_negative | Non_nature | Total |
| --- | --- | --- | --- | --- | --- | --- | --- | --- | --- | --- | --- |
| Green-Activity |  |  |  |  |  | Blue-Activity |  |  |  |  |  |
| run | 106 | 89 | 34 | 650 | 879 | swim | 19 | 36 | 20 | 0 | 75 |
| walk | 123 | 138 | 61 | 20 | 342 | dive | 3 | 3 | 2 | 64 | 72 |
| bicycle | 60 | 93 | 15 | 31 | 199 | boat | 5 | 3 | 2 | 33 | 43 |
| football | 7 | 62 | 8 | 46 | 123 | surf | 2 | 2 | 1 | 22 | 27 |
| hike | 50 | 43 | 8 | 7 | 108 | fishing | 3 | 6 | 0 | 13 | 22 |
| Green-Place |  |  |  |  |  | Blue-Place |  |  |  |  |  |
| outside | 81 | 186 | 53 | 311 | 631 | beach | 24 | 62 | 21 | 23 | 130 |
| park | 61 | 93 | 21 | 52 | 227 | pool | 7 | 24 | 16 | 44 | 91 |
| sun | 27 | 43 | 10 | 76 | 156 | water | 3 | 2 | 1 | 75 | 81 |
| nature | 32 | 15 | 2 | 84 | 133 | sea | 2 | 7 | 1 | 24 | 34 |
| camp | 14 | 45 | 8 | 38 | 105 | stream | 1 | 0 | 0 | 32 | 33 |

### 3.2 Topic Modeling

The BERTopic model was created from the 1,887 nature-related posts. After paragraph segmentation, there were 7,436 documents used in the model. The model identified 14 distinct topics in the data (Figure 4). Topics labels were generated by human interpretation and automatically using state-of-the-art LLMs, ChatGPT-4o and Claude Sonnet 3.7, with the final label determined by consensus. The prompt and the result from each model can be found in Supplemental Material S2.

**Figure 4:**
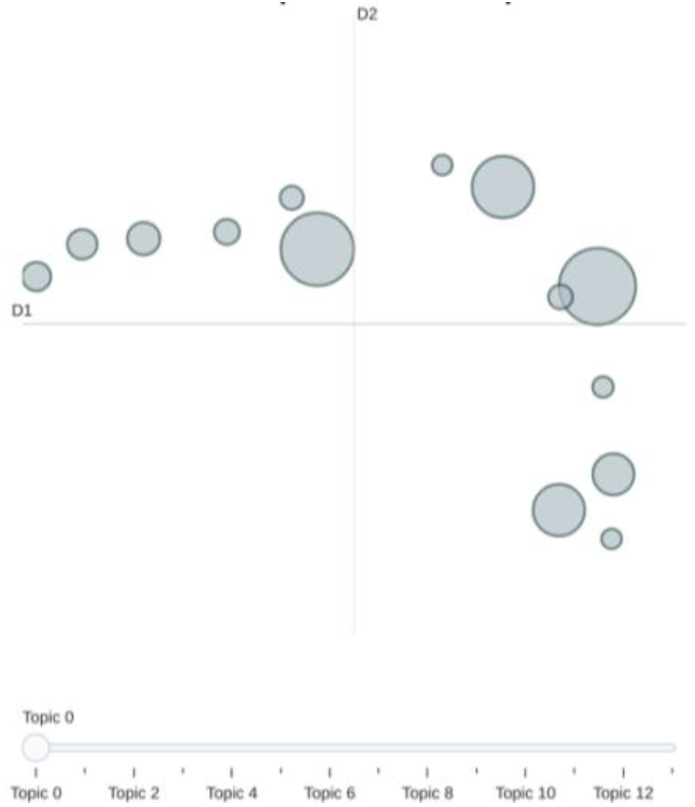
The intertopic distance map shows the relationship between the topics; the closer the center of the circle, the more related the topics are. The map shows the distinctiveness of the generated topics. The size of the circle represents the number of documents in the topic.

The labeled 14 topics with their keyword representation and some example texts are found in Table 5. BERTopic puts outlier documents, or noise, that do not fit into the other topics into a separate topic (−1). Our model classified 26.6% (1977/7436) of documents as outliers.

**Table 5:**
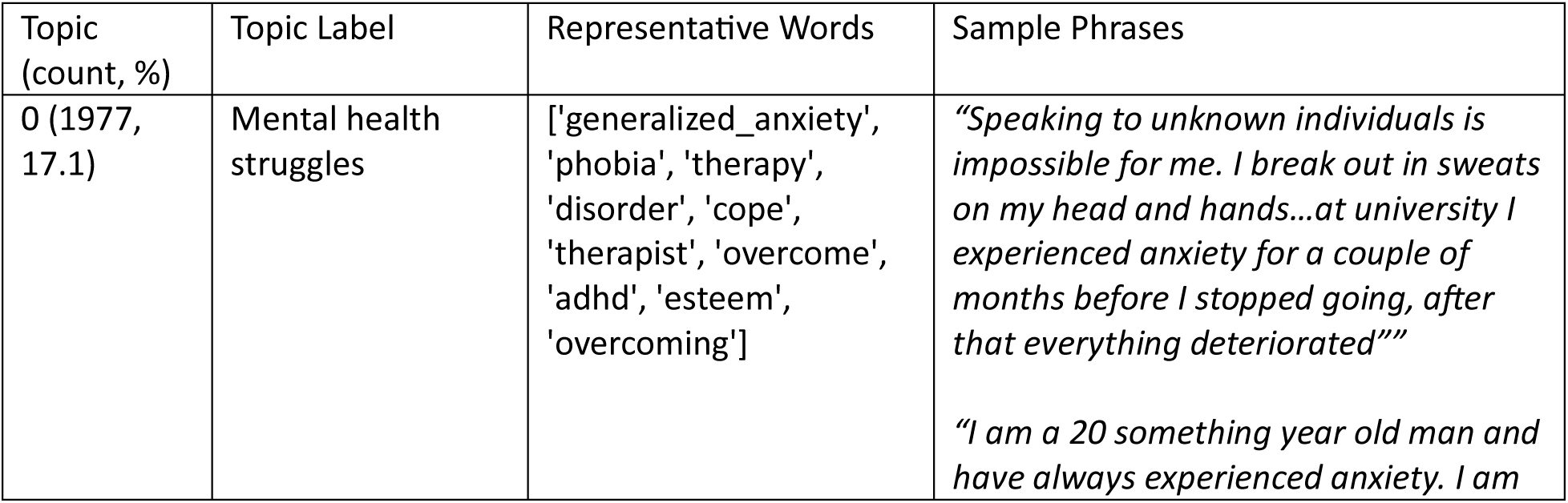
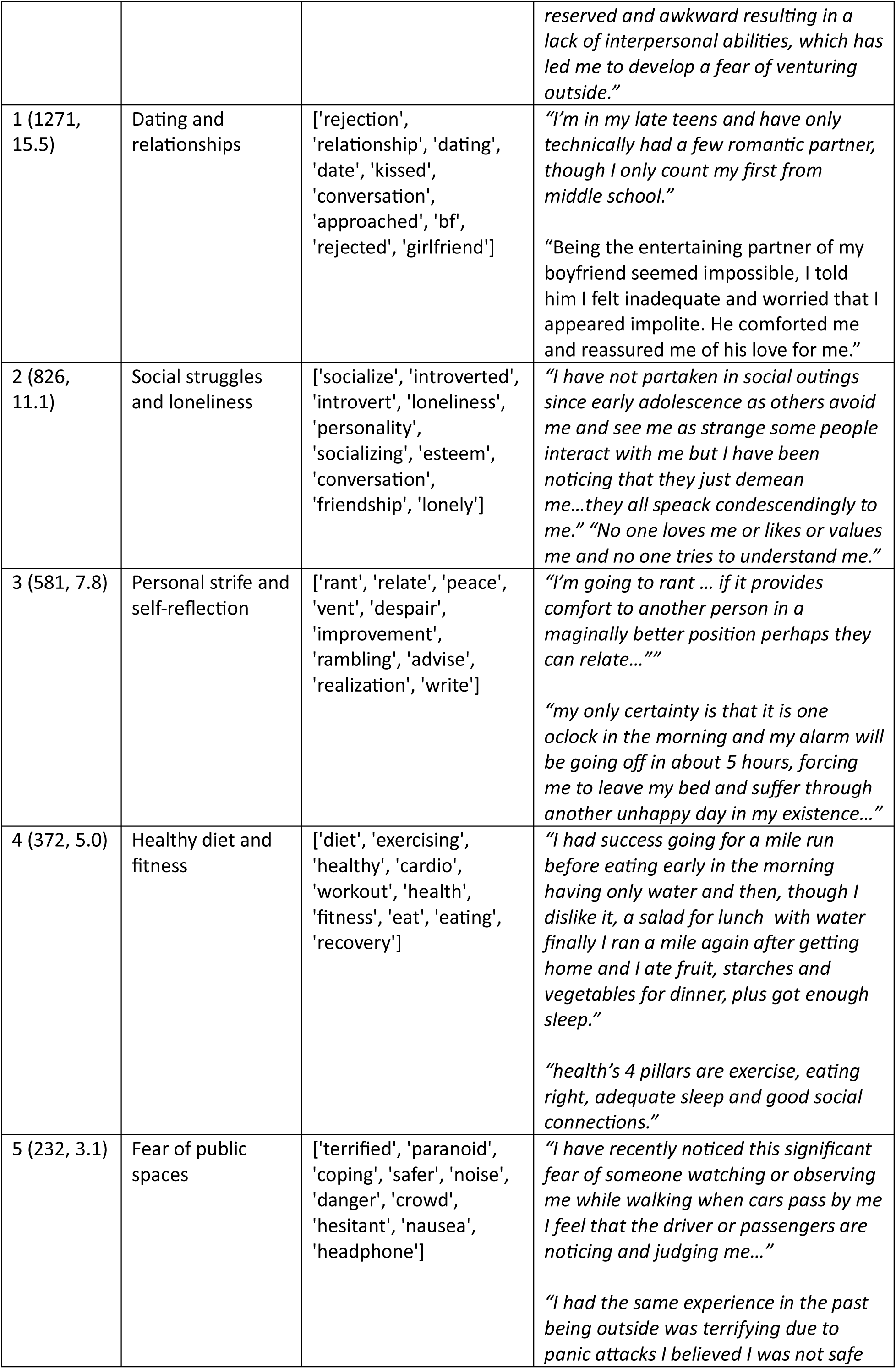
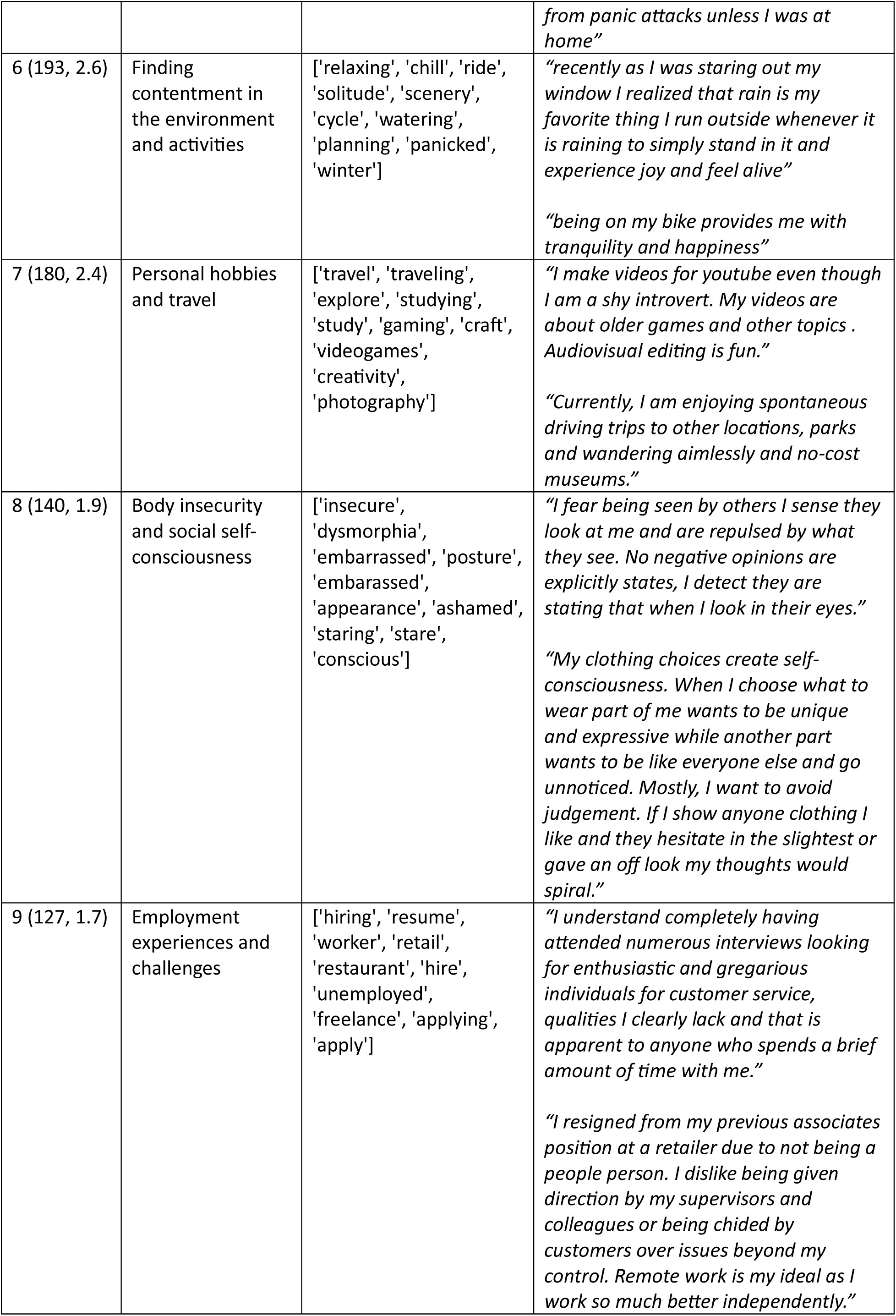
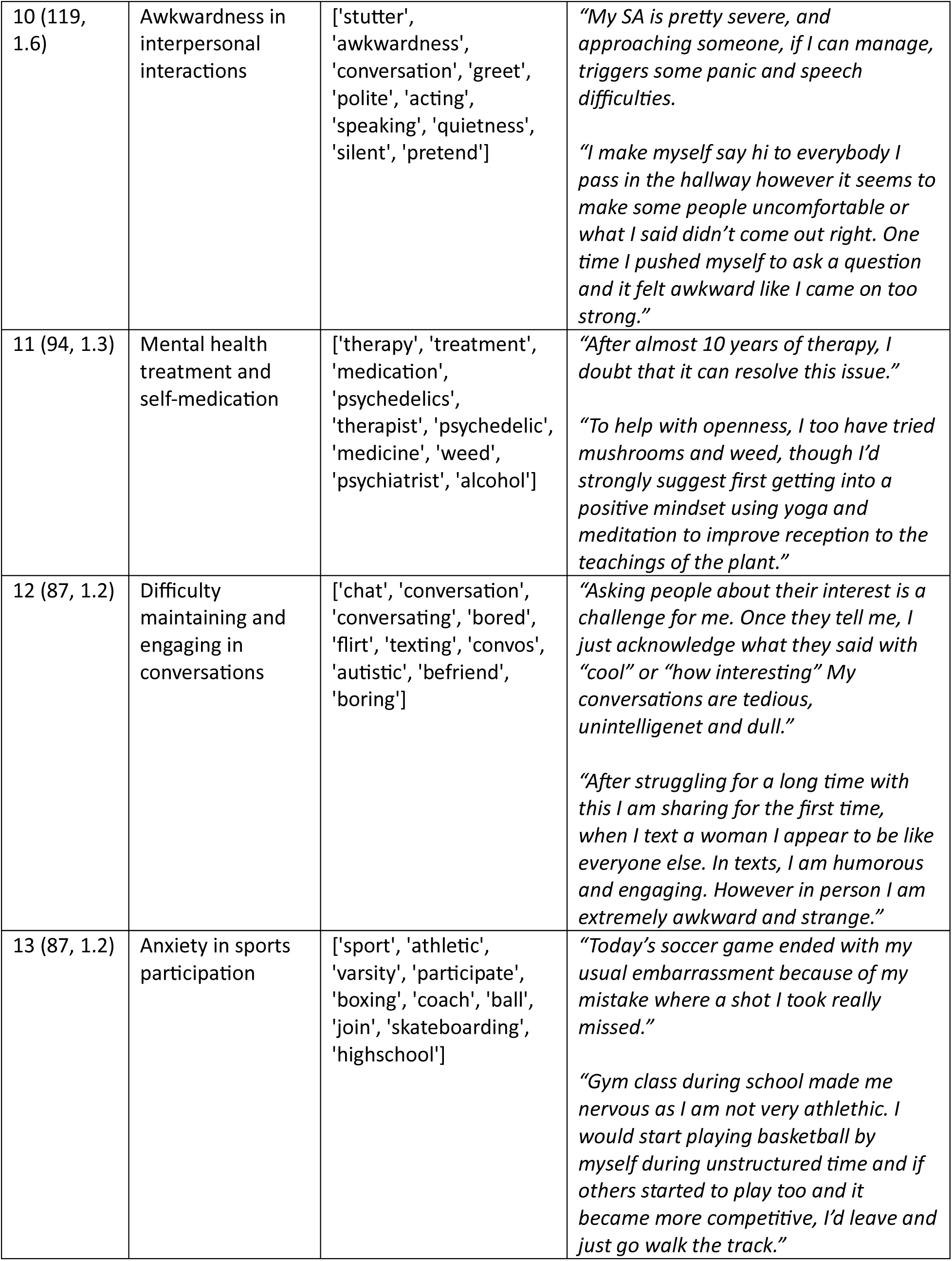
14 topics identified by the BERTopic model with their topic label, keyword representation, and example phrases from the paragraph.

The 5,459 paragraph segments that were assigned a topic came from 1,669 posts. The number of unique topics per post ranged from 1 to 7, with an average of 1.87 unique topics per post. About half of the posts, 50.9% (848/1,669), had only one topic assigned, while 3.4% (57/1,669) had five or more topics assigned.

Several similarities and differences can be observed from the co-occurrence network of topics in the positive and negative class posts (Figure 5). Both classifications show strong connections between mental health struggles (topic 0) and personal strife and self-reflection (topic 3). Social struggles and loneliness (topic 2) are also highly connected in both classes. Positive posts have stronger connections to a healthy diet and fitness (topic 4), personal hobbies and travel (topic 7), and employment experiences and challenges (topic 9). While fear of public spaces (topic 5) and body insecurity and social self-consciousness (topic 8) were more prevalent in negative posts. Negative posts have fewer unique topic connections overall (15 vs. 19 connections).

**Figure 5:**
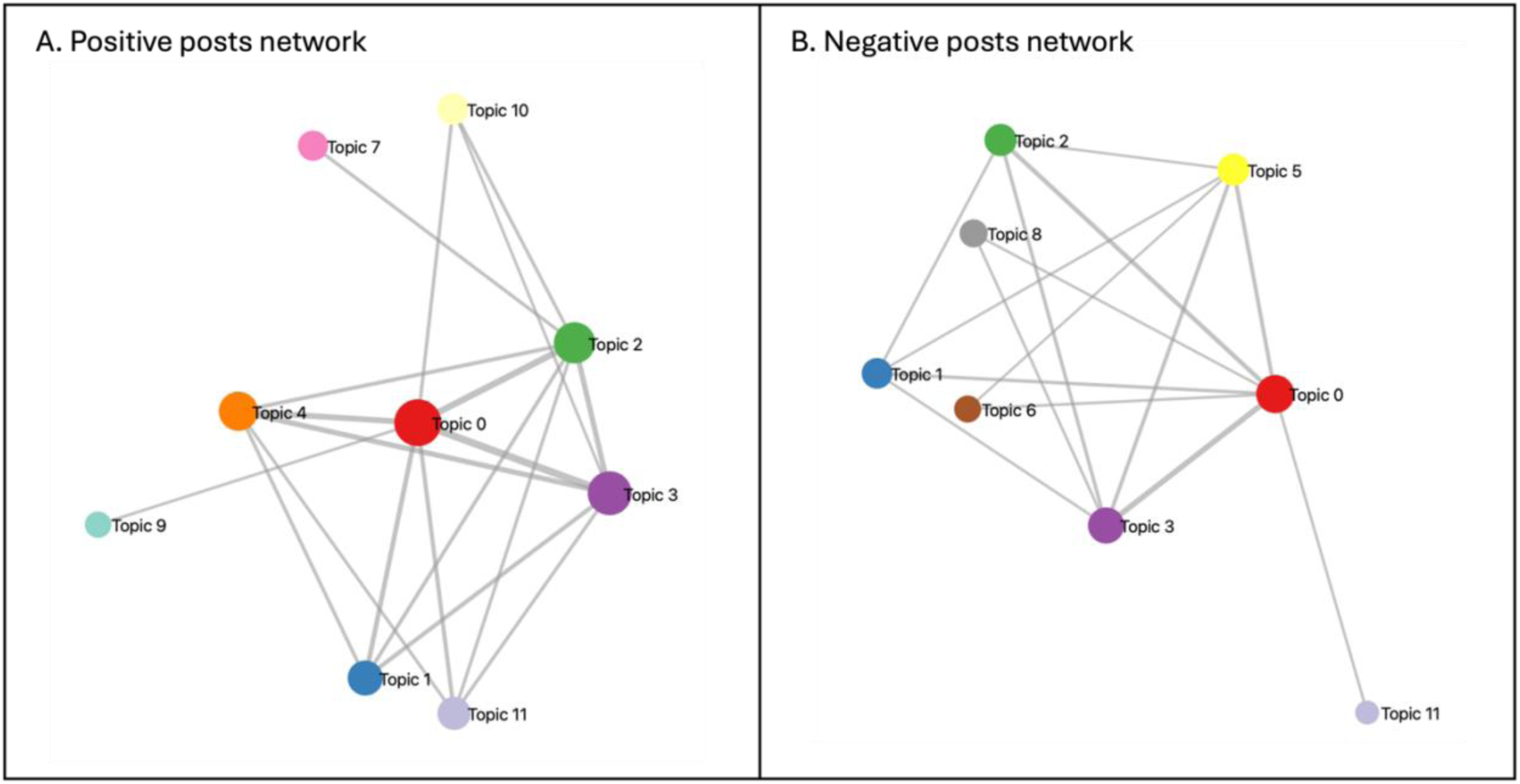
Co-occurrence networks for positive (panel A) and negative (panel B) posts. Node size indicates frequency, and link thickness represents co-occurrence strength.

### 3.3 Qualitative Analysis

A total of 307 positive and negative posts were manually coded in NVivo. Within the six themes, 378 phrases were coded with their underlying topics. For this analysis, topics were only counted once per post. The total counts by theme and code, including the total number of references coded, are in the supplemental material (S3). Of all the themes, only one encompassed the negative perceived impact of the natural environment on the user’s SAD. This theme, ‘challenges and barriers’, had 100 coded phrases (26.5%, 100/378). The remaining five themes reflected a positive perceived impact on the user’s well-being. Of these themes, ‘positive impact on wellbeing’ had the most phrases identified with 126 (33.3%, 126/378) The remaining themes were ‘advice to others’ with 65 phrases (17.2%, 65/378), ‘coping strategies’ (12.2%, 46/378), ‘enhanced social connections’ (6.6%, 25/378) and ‘restorative environment’ (4.2%, 16/378)

#### 3.3.1 Challenges and barriers (n= 100)

This theme included five topics, covering the user’s negative perceptions of being in the natural environment or an inability to access these environments. Three topics related to the user’s stated mental and physical well-being. Discussions in two of the topics, ‘triggers stressor’ (60%, 60/100), ‘causes symptoms (15%, 15/100) centered on how being in the natural environment or participating in activities in these spaces causes, or they fear it will cause them to experience physical or mental symptoms of their SA, or discomfort in general.

> *“I stopped attending horse riding lessons due to unreasonable beliefs, such as “people dislike me ..I create problems when I show up” despite everyone being kind and supportive.”*
>
> *“…anxiety symptoms when leaving home particularly at a crowded location such as the beach”*

The third topic, ‘unable to due to SA’ (16% 16/100) was similar, however, in these the user explicitly stated that they could not bring themselves to be in or participate in activities due to their SA.

> *“Running is a favorite sport of mine, yet am overwhelmed by the anxiety about encountering another person on my side of the street or someone waving at me.”*

The remaining topics in this theme were related to users discussing the natural environment not being available to them (‘unavailable’, 5%, 5/100) or not accessible (‘no access’, 4%, 4/100) for them.

> *“Due to my current financial situation, I cannot afford to go to the lake”*

#### 3.3.2 Positive impact on well-being (n= 126)

This theme included four topics related to the user expressing that they perceived that the natural environment had beneficial effects on their mental and physical health. In the topic ‘positive emotion’ (60.3%, 76/126), the user expressed feeling good or having an improvement in mood by being in the natural environment. While the topic ‘alleviates symptoms’ (5.6%, 7/126) expressed similar sentiment, but for more physical symptoms.

> *“I was with family and spent time outdoors, enjoying focusing on myself, not worrying about other people and appreciating the scenery.”*
>
> *“My anxiety would probably be minimal. Perhaps others experience this too, but being outside in the fresh, cooler air and in more open spaces mimimizes blushing and helps me stay composed.”*

For the topic ‘calming and relaxing’ (16.7%, 21/126), the user specified that they find being in or using the natural environment or activity to calm or relax them.

> “taking a walk outside in nature with music relaxes me every time”

The final topic in the theme, ‘wish to use it to help’ (17.5%, 22/126), was used to code expressions by the user, who relayed a sense that they felt the natural environment would help or be beneficial to them to alleviate their SA.

> “Activities I’ve considered trying are solo camping or backpacking trips.”

#### 3.3.3 Advice to others (n=65)

This topic, related to this theme, was ‘encourages action’. This occurred in posts where the user was replying to another poster in the subreddit. These posts encouraged the poster to use the natural environment to help them overcome their SA or the symptoms of their SA. The user either stated explicitly that these environments or activities had helped them, or it was implied in their post.

> *“Find a productive outlet for the anxious feelings, such as positive diversions like the gym, running, swimming or something else you enjoy, daily after work.”*

#### 3.3.4 Coping strategies (n=46)

This theme included two topics: ‘exposure therapy’ (56.5%, 26/46) and ‘facilitates behavior change’ (43.5%, 20/46). For both these topics, the user expressed that they used the natural environment or activities to help them overcome their SA symptoms or improve the condition itself.

> *“My plan tomorrow is to ride the bike trail nearby with a goal of greeting five others on the trail. This is an action I am implementing to reduce some of my SA symptoms.”*
>
> *“At the end of the semester, I’m going backpacking overseas. Encountering strangers who won’t know of my SA struggles at home.”*

#### 3.3.5 Enhanced social connection (n=25)

This theme centered on users discussed how the natural environment facilitated their connections to others and thus helped manage or overcome their SA. In the topic ‘connection to others’ (60%, 15/25), users discuss how they were able to enjoy the company of others while being in the natural environment. This allowed them to overcome some of the fears they experienced with their SA.

> *“I sat on a bench and spoke with someone I didn’t know for ten minutes…she sat next to me and explained she was waiting for someone too, we exchanged pleasantries..it was an enjoyable encounter.”*

The ‘shared interest’ (40%, 10/25) topic consisted of discussions around how having a shared interest in outdoor environments or activities with others facilitated the user’s interaction with them in ways they may have struggled with in other situations due to their SA.

> *“For instance, we were guiding kayakers….so I wasn’t alone but was supervising. Because I had a purpose, I found conversation and activities easier and I was less introverted.”*

#### 3.3.6 Restorative environment (n=16)

The final two topics are under the theme of restorative environment. The topic ‘reduces stress’ (62.5%, 10/16) was encoded for mentions of the natural environment in conjunction with the user feeling a reduction in stress or anxiety.

> “To help develop self-awareness during stressful situations, I used a mindfulness practice app. I would sit on a bench at a local park to practice with it.”

While in ‘restores attention’ (37.5%, 6/10) posts, users discussed how the natural environment helps to relieve their stress or anxiety.

> *“I was unable to deal with the inactivity at my home, so I took my sibling’s unused longboard, some earbuds and music and went out with no particular destination. My head cleared immensely with this activity.”*

## 4. Discussion

The purpose of this preliminary study is to explore the relationship between natural environments and their potential effects on social anxiety disorder by analyzing the perspectives of adolescents and young adults on Reddit. Our findings revealed a complex relationship between SAD and engagement with green and blue spaces, with both positive and negative impacts identified.

Our findings that outdoor environments and activities had predominantly positive effects on SAD symptoms are consistent with prior research showing that exposure to green space is linked to lower anxiety ^38^, including benefits for young adults^39^. However, our analysis extends previous work by highlighting the unique challenges that individuals with SAD face when engaging with natural environments. Unlike those with generalized anxiety disorder (GAD), individuals with SAD will experience anxiety specifically in social situations where they may be judged or scrutinized ^4^ . Indeed, we did observe in our cohort that SAD sufferers described specific fears related to social evaluation in outdoor settings. This finding suggests that nature-based interventions for SAD may need tailoring to address social evaluation concerns, rather than simply replicating approaches used for GAD.

The finding that participants often use nature as a form of exposure therapy reflects clinical recommendations for SAD treatment. Meta-analyses of SAD treatments have shown that exposure therapy is among the most effective interventions ^11,40^, and our findings suggest that natural environments may provide a less threatening context for practicing social interactions compared to more formal clinical settings or smaller, indoor settings, where the user’s fear of judgment may be heightened.

Our analysis revealed that green environment activities (such as walking, running, and cycling) were most frequently associated with positive impacts, consistent with research showing that physical activity in natural settings provides greater mental health benefits than indoor exercise ^41,42^. This effect of nature exposure and physical activity may be particularly beneficial for SAD, as it addresses both psychological symptoms and the physical manifestations of anxiety.

Our results indicate that the perception of the natural environment from users with SAD aligns with the theoretical frameworks for the cognitive benefits of natural environments ^43^ . The qualitative analysis identified six key themes corresponding to existing nature and mental health theories. The “restorative environment” theme, which included codes like “reduces stress” and “restores attention,” directly supports ART ^22^. As proposed by ART, participants described how natural environments provided cognitive restoration, supporting the theoretical mechanism that natural settings help replenish depleted cognitive resources. For example, participants reported that activities like biking while listening to music helped “clear their head,” consistent with ART’s proposition that natural environments facilitate cognitive recovery.

Similarly, the “positive impact on well-being” theme, particularly codes like “calming and relaxing,” aligns with SRT ^24^. Participants frequently described feeling relaxed and experiencing reduced physiological stress responses when in natural settings, supporting SRT’s hypothesis that natural environments trigger parasympathetic responses that reduce physiological stress markers.

Our analysis’s “enhanced social connection” theme provides empirical support for the Biophilia Hypothesis ^25^. Participants reported that natural environments facilitated social connections through shared interests and activities, suggesting that these settings may offer opportunities for connection without the intimidating evaluation concerns typically experienced in other social contexts. These results align with the recent understanding of the role of nature on mental health from the emerging field of environmental neuroscience ^44,45^.

Our mixed-methods approach, combining computational topic modeling with in-depth qualitative analysis, provided both breadth and depth in understanding SAD experiences with natural environments. The large-scale topic modeling identified patterns across thousands of posts, providing high-level insights into the patients’ perspectives on the influence of the natural environment on their SAD. However, such analysis may miss relevant context or nuances in the posts. However, our qualitative analysis filled these gaps by providing rich contextual insights into the text underlying these patterns. Additionally, our focus on adolescents and young adults (13-25 years) captures a critical developmental period for SAD onset and manifestation, providing valuable insights into how natural environments may affect this vulnerable population during a formative life stage.

Using Reddit as a data source allowed unobtrusive access to discussions from a population that is typically difficult to reach through traditional research methods. Individuals with SAD often avoid research participation due to social evaluation concerns, making online platforms valuable for capturing authentic experiences without the factors that limit the appeal of formal research settings for this population.

However, some limitations should be considered when interpreting our findings. The self-reported nature of Reddit posts means we cannot verify SAD diagnoses. Users might be discussing self-diagnosed symptoms rather than clinically diagnosed SAD, potentially limiting generalizability to clinical populations. As with any research using data from social media, our data may not fully represent the population at large. While the Reddit users in our cohort were selected based on a predefined age range, other demographic information about them is unknown, thus limiting the generalizability of our findings to the population at large.

Finally, while our keyword filtering approach was comprehensive, it may have missed relevant discussions that used alternative terminology to describe natural environments or SAD experiences. Future research could employ more sophisticated natural language processing techniques to identify relevant content without relying on predefined keywords or regular expression patterns.

### Privacy and Ethics Statement

This study was reviewed by the Institutional Review Board of the University of Pennsylvania and Cedars-Sinai Medical Center, and deemed exempt from human subjects research under Category (4) of Paragraph (b) of the US Code of Federal Regulations Title 45 Section 46 for publically available data sources. The study is an observational study involving the analysis of publically available Reddit data with no direct interaction or intervention with users. All procedures followed institutional guidelines and adhered to the British Psychological Society^46^, the Association of Internet Researcher (AoIR)^47^ guidelines and recommendations for internet research. The data was collected in adherence to Reddit’s Terms of Service and PushShift’s API usage policies in effect at the time of data collection (March 2022).

Reddit allows users to post under pseudonyms, limiting the ability to link posts to a particular person. To further preserve the privacy of the users, all example quotes from Reddit posts presented in this manuscript have been modified to prevent re-identification through internet searches. These modifications preserve the semantic meaning and all nature-related keywords which were essential to the study’s analysis. Additionally, all findings are reported in aggregate and no user names are reported.

## 5. Recommendations

Based on the results of this research, recommendations can be made to address the increasing rates of social anxiety ^48^. Firstly, it has been found that spending 120 minutes in a natural environment results in good health and well-being ^49^. Given this and the results of this paper, outdoor activities and time outdoors may be considered as potentially beneficial for alleviating social anxiety symptoms ^50^.

Second, given the study’s findings that walking, running, and biking outdoors significantly enhance positive emotions, it is recommended that urban areas with high populations incorporate multiple green walking routes to parks to help alleviate social anxiety symptoms. Residents residing in cities compared to rural environments were found to be 21% more likely to have anxiety disorders ^51^. Cities and urban areas are also less green than rural areas, resulting in less connection to green areas ^52^. Within cities, blocks with more than 10 trees decrease hypertension risk ^53^. This reasoning also supports the third recommendation of expanding existing parks and increasing the number of parks that more accurately reflect an area’s population. 1 in 3 U.S. residents do not have a park or green space within a 10-minute walk of their homes, translating to 100 million people not having walkable access to parks ^54^. To accomplish this recommendation, cities need to assess current park distribution and access, identify areas lacking park access, consider population density in underserved areas, evaluate available land for park development, and develop and increase parks and park access with geographical and urban planning constraints.

## 6. Conclusion

This study provides novel insights into how individuals with SAD experience and engage with natural environments. Our findings suggest that despite challenges related to social evaluation concerns, many individuals with SAD find natural spaces beneficial for symptom management, cognitive restoration, and facilitating positive social interactions. The therapeutic potential of natural environments aligns with established theoretical frameworks of nature’s restorative properties and existing clinical approaches for SAD treatment.

While many users discuss the benefits of these natural environments, others reported fear of, or the exacerbation of symptoms, in these places. These differences in experiences identified highlight the importance of personalized approaches to nature-based interventions for SAD. Simply recommending outdoor activity may be insufficient for this population; however, thoughtfully designed nature experiences that manage social evaluation concerns while leveraging the restorative aspects of natural environments could provide a valuable complementary approach to traditional SAD treatments. Community planners may want to cognitively design inclusive spaces for people with SAD.

As urbanization increases and mental health concerns like SAD continue to rise, particularly among young people, integrating accessible natural environments into community planning and healthcare approaches represents an evidence-informed strategy that could improve outcomes and the quality of life for individuals with SAD while promoting public health more broadly.

## Data Availability

The data underlying this study were obtained from publicly available Reddit posts and comments and are subject to Reddit’s Public Content Policy and terms of use. The authors are not permitted to redistribute the original Reddit content or any dataset containing it. Analysis of Reddit data of this type is allowed, but each researcher must independently collect the underlying data from Reddit in accordance with Reddit’s terms and any applicable institutional ethics requirements. All annotation guidelines, code, and detailed instructions necessary to reproduce the data collection and analysis are reported in this manuscript and openly available at **10.5281/zenodo.18331364** .

## Author Contribution

KO participated in the experimental design, perfomed experiments, developed coding schema, analyzed data, drafted and edited the manuscript, and created figures, SH participated in conception of the study, developmed coding schema, analyzed data and interpreted results, drafted the manuscript, ALS procured and filtered the data, contributed to the development of the manuscript, PS participated in designing the study and edited the manuscript, GGH participated in the conception and desgn of study, edited the manuscript and secured funding for the study.

## Funding Declaration

This work was supported by the National Institutes of Health (NIH) National Library of Medicine (NLM) under grant NIH-NLM R01LM011176. The NIH NLM funded this research but was not involved in the design or conduct of the study; collection, management, analysis, or interpretation of the data; preparation, review, or approval of the manuscript; or the decision to submit the manuscript for publication.

---

**Table S1:**
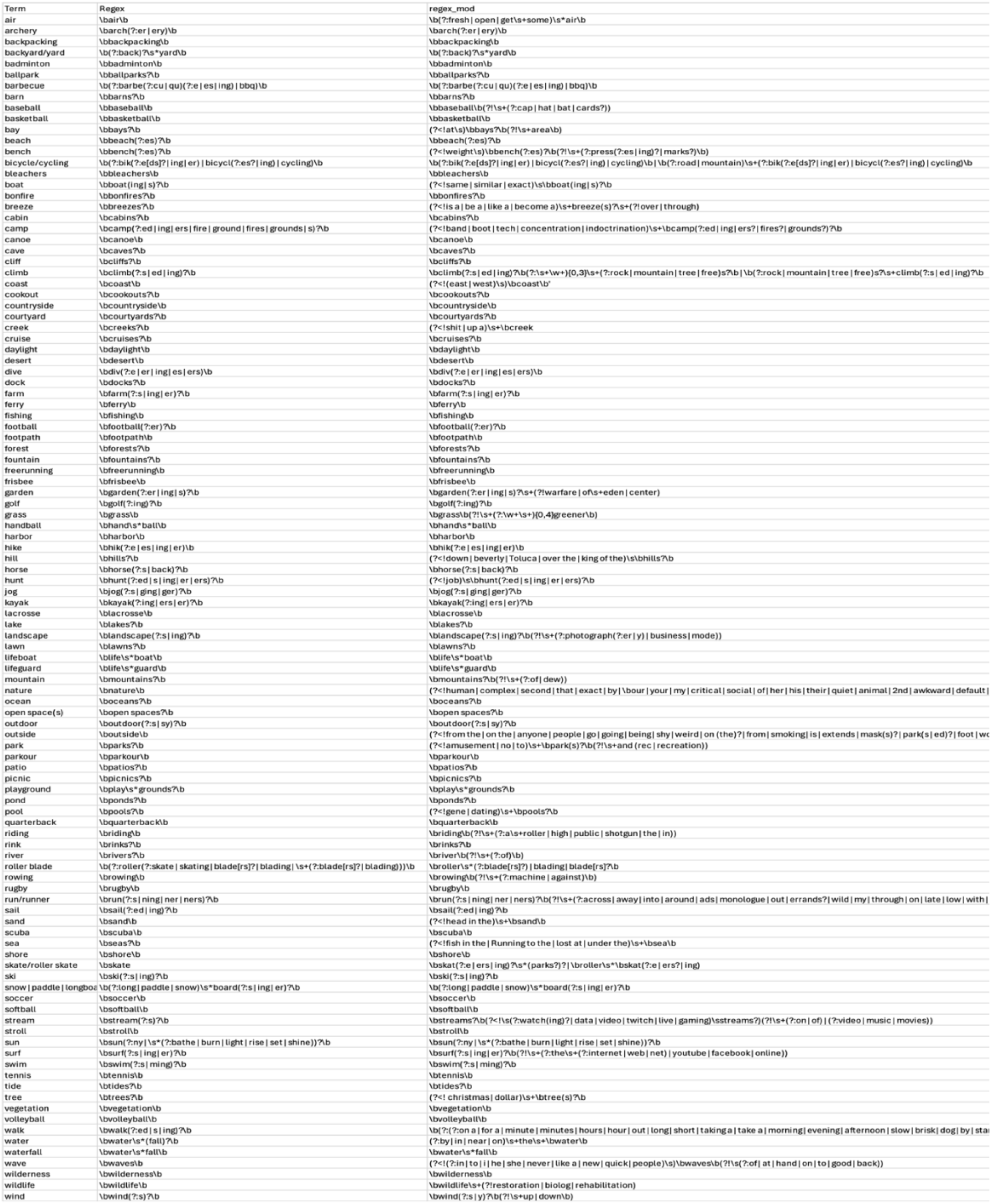
Keywords and regular expressions.

**Table S2:** Prompt used for the assignment of labels by the LLM and the results. Prompt: “*I have a topic that contains the following documents: [documents in attachment} The topic is described by the following keywords [list of representative keywords]* *Based on the information above, extract a short topic label in the following format: topic:* “

| Topic | KeybertInspired Representations | Human annotator | Chat-GPT 4o | Claude Sonnet 3.7 | Final Topic Label |
| --- | --- | --- | --- | --- | --- |
| 1 | 'therapy', 'disorder', 'cope', 'therapist', 'overcome', 'adhd', 'esteem', 'overcoming'] | mental health issues | Social anxiety and mental health struggles | Anxiety disorders and mental health struggles | Mental health struggles |
| 2 | 'date', 'kissed', 'conversation', 'approached', 'bf', 'rejected', 'girlfriend'] | dating/romantic relationships | Dating, relationships and social anxiety | Dating and relationship anxiety | Dating and relationships |
| 3 | ['socialize', 'introverted', 'introvert', 'loneliness', 'personality', 'socializing', 'esteem', 'conversation', 'friendship', 'lonely'] | Loneliness/difficulty socializing | Social struggles and introversion | Social anxiety and loneliness | Social struggles and loneliness |
| 4 | ['rant', 'relate', 'peace', 'vent', 'despair', 'improvement', 'rambling', 'advise', 'realization', 'write'] | Asking for advice or venting | Personal struggles and self-reflection | Personal struggles and self-reflection | Personal strife and self-reflection |
| 5 | ['diet', 'exercising', 'healthy', 'cardio', 'workout', 'health', 'fitness', 'eat', 'eating', 'recovery'] | diet and exercise | Healthy diet and fitness | Personal health and fitness improvement | Healthy diet and fitness |
| 6 | ['terrified', 'paranoid', 'coping', 'safer', 'noise', 'danger', 'crowd', 'hesitant', 'nausea', 'headphone'] | fear of outdoor spaces | Social anxiety and public distress | Social anxiety and fear of public spaces | Fear of public spaces |
| 7 | ['relaxing', 'chill', 'ride', 'solitude', 'scenery', 'cycle', 'watering', 'planning', 'panicked', 'winter'] | relaxing environments/activities | Anxiety and solitude in everyday experiences | Social anxiety and finding peace | Finding contentment in the environment and activities |
| 8 | 'studying', 'study', 'gaming', 'craft', 'videogames', 'creativity', 'photography'] | hobbies | Creativity, travel and exploration | Travel, creating and personal hobbies | Personal hobbies and travel |
| 9 | 'embarrassed', 'posture', 'embarrassed', 'appearance', 'ashamed', 'staring', 'stare', | self-judgement/insecurities | Body image anxiety and social self consciousness | Body image insecurities and social anxiety | Body insecurity and social self consciousness |
| 10 | ['hiring', 'resume', 'worker', 'retail', 'restaurant', 'hire', 'unemployed', 'freelance', 'applying', 'apply'] | employment | Employment challenges and job search | Social anxiety and employment challenges | Employment experiences and challenges |
| 11 | 'conversation', 'greet', 'polite', 'acting', 'speaking', 'quietness', 'silent', 'pretend'] | awkwardness socializing in person | Social anxiety and awkwardness in conversation | Social anxiety and face-to-face interactions | Awkwardness in interpersonal interactions |
| 12 | 'psychedelics', 'therapist', 'psychedelic', 'medicine', 'weed', 'psychiatrist', 'alcohol'] | treatments for SA | Mental health treatment and substance use | Mental health treatment and self-medication | Mental health treatment and self-medication |
| 13 | 'conversating', 'bored', 'flirt', 'texting', 'convos', 'autistic', 'befriend', 'boring'] | inability to maintain conversations | Struggles with conversation and social interaction | Difficulty maintaining and engaging in conversations | Difficulty maintaining and engaging in conversations |
| 14 | ['sport', 'athletic', 'varsity', 'participate', 'boxing', 'coach', 'ball', 'join', 'skateboarding', 'highschool'] | participating in team sports | Struggles and anxiety in sports participation | Negative experience and anxiety in youth sports participation | Anxiety in sports participation |

**Table S3:** Counts of the final coding from the qualitative analysis performed using NVivo.

| Code Name | Unique Count | Total References |
| --- | --- | --- |
| Advice to others |  |  |
| Encourages action | 65 | 70 |
| Challenges and Barriers |  |  |
| Causes Symptoms | 15 | 16 |
| No access | 4 | 4 |
| Triggers stressors | 60 | 70 |
| Unable due to SA | 16 | 16 |
| Unavailable | 5 | 5 |
| Coping Strategies |  |  |
| Exposure Therapy | 20 | 20 |
| Facilitates behavior change | 26 | 26 |
| Enhanced Social Connection |  |  |
| Connection with others | 15 | 16 |
| Shared interest | 10 | 10 |
| Positive impact on wellbeing |  |  |
| Alleviates symptoms | 7 | 7 |
| Calming and relaxing | 21 | 21 |
| Positive emotion | 76 | 81 |
| Wish to use it to help | 22 | 22 |
| Restorative Environment |  |  |
| Reduces stress | 10 | 10 |
| Restores attention | 6 | 6 |
| Grand Total | 378 | 400 |

## Notes

### Competing Interest Statement

The authors have declared no competing interest.

### Author Declarations

Reddit downloaded from Pushshift.io

### Summary of Updates

Updated privacy and ethics statement, and data availability statement

